# Effectiveness, safety and acceptability of no-test medical abortion provided via telemedicine: a national cohort study

**DOI:** 10.1101/2020.12.06.20244921

**Authors:** Abigail Aiken, Patricia A Lohr, Jonathan Lord, Nabanita Ghosh, Jennifer Starling

## Abstract

**Objectives:** To compare the effectiveness, safety and acceptability of medical abortion before and after the introduction of no-test telemedicine abortion care.

**Design:** Cohort study to assess whether a no-test telemedicine-hybrid care model (telemedicine with in-person provision only when indicated) was non-inferior to the traditional service model (blanket in-person provision including ultrasound scan).

**Setting:** The three main abortion providers in England and Wales.

**Participants:** All patients having an early medical abortion in the two months before and after the service model change. Patient demographic and clinical characteristics were compared between the cohorts to adjust for any systematic differences in the two groups.

**Main outcome measures:** *Access:* waiting time, gestation at abortion

*Effectiveness:* the proportion of successful medical abortions

*Safety:* significant adverse events defined as: haemorrhage requiring transfusion, significant infection requiring hospital admission, major surgery, death. We also examined the incidence of ectopic pregnancy and late gestation.

*Acceptability:* Patient-reported outcomes of satisfaction, future preference, and privacy of consultation

**Results:** The study sample included 52,142 medical abortions; 22,158 in the traditional cohort and 29,984 in the telemedicine-hybrid cohort, of which 61% were provided using no-test telemedicine. The cohorts accounted for 85% of all medical abortions provided in England and Wales during the study period. Mean waiting times were 4.2 days shorter in the telemedicine-hybrid cohort, and 40% were provided at ≤6 weeks’ gestation compared to 25% in the traditional cohort (p<0.001). There was no difference in success rates between the two groups (98.2% vs. 98.8%, p=1.0), nor in the prevalence of serious adverse events (0.04% vs. 0.02%, p=0.557). The incidence of ectopic pregnancy was equivalent in both cohorts (0.2%, p=0.796), with no significant difference in the proportions being treated after abortion (0.01% vs 0.03%, p=0.123). In 0.04% of cases the abortion appeared to have been provided at over 10 weeks’ gestation; these abortions were all completed at home without additional medical complications. In the telemedicine-hybrid group, the effectiveness for abortions conducted using telemedicine (n=18,435) was higher than for those conducted in-person (n=11,549), 99.2% vs. 98.1%, p<0.001. Acceptability was high (96% satisfied), 80% reported a future preference for telemedicine and none reported that they were unable to consult in private using teleconsultation.

**Conclusions:** Medical abortion provided through a hybrid model that includes no-test telemedicine without routine ultrasound is effective, safe, acceptable, and improves access to care.

**Summary Box:** 

**What is already known on this topic:** The UK’s National Institute for Health and Care Excellence (NICE) conducted a systematic review and recommended using telemedicine to improve access to medical abortion care.

Several models for using telemedicine to facilitate medical abortion have been described, but most existing trials are small, and many required attendances to have medicines administered or for an ultrasound scan or blood tests.

**What this study adds:** This study (n=52,142) is the first to assess a real-world no-test telemedicine abortion care pathway in a national population. The new national model demonstrates how a permissive framework for medical abortion can deliver significant quality improvements to those needing to access abortion care. There was no difference in effectiveness (p=1.0) or safety (p=0.6) when compared to a traditional in-person model, but the no-test telemedicine pathway improved access to care, was highly acceptable to patients and is likely to be especially beneficial for vulnerable groups and in resource-poor settings.

## Introduction

Improved access to abortion care would deliver significant advantages for both healthcare systems and the women who use them. Abortion is a common reason for needing healthcare – the global abortion rate is estimated at 39 abortions per 1000 women aged 15–49 years,^1^ with 28% of all pregnancies in developed countries ending in abortion.^2^ There is clear evidence that restricting access to abortion does not reduce abortion rates, it simply makes abortion less safe.^1 3^ Improving access is likely to benefit those who are most vulnerable,^4^ especially in resource-poor settings or where care has to be self-funded. In its 2019 guideline on abortion care, the National Institute for Health and Care Excellence (NICE) stated that improving access to abortion was a key priority.^5^

Telemedicine, the use of information and communication technologies to improve patient outcomes by increasing access to care and medical information,^6^ has been noted to decrease costs and increase convenience and safety.^7^ It is an established service delivery model for abortion care in many settings,^8^ and it is recommended to improve access.^4^ The COVID-19 pandemic required urgent action by clinicians and policymakers to ensure delivery of essential health services, including abortion. In March 2020, the Royal College of Obstetricians and Gynaecologists (RCOG) published guidelines to safeguard abortion care in the UK.^9^ The guidelines profoundly changed the way medical abortion care is delivered in Great Britain. Prior to the emergence of COVID-19, all patients seeking medical abortion were required to attend in-person to receive an ultrasound scan and have mifepristone administered within the clinic. Under the new guidelines, consultations were encouraged to take place by telephone or video call; an ultrasound scan was required only if indicated. All the British governments had issued emergency legal orders by March 30^th^ 2020 to allow mifepristone to be used at home along with misoprostol up to 10 weeks’ gestation.^10-12^ These approvals permitted abortion providers to implement a telemedical service delivery model for medical abortion.

Great Britain’s new pathway for no-test telemedicine abortion is unusual among existing models because it is fully remote: no clinic visit, tests or ultrasound scan are required and both mifepristone and misoprostol are delivered by mail or collected from a clinic for use at home. This new service model thus presents an important opportunity to evaluate a potentially better way to provide medical abortion that could improve access and reduce the barriers posed by in-person care.^13^ We examined and compared the effectiveness, safety and acceptability of medical abortion provided up to 10 weeks’ gestation before and after the service model change.

## Methods

### Population and Cohorts

The study population comprised all patients who accessed an early medical abortion (EMA) at the three largest abortion providers in England and Wales - British Pregnancy Advisory Service (BPAS), MSI Reproductive Choices (MSUK), and the National Unplanned Pregnancy Advisory Service (NUPAS) - two months before and after the service model change. Medical abortion is defined as the use of medications to cause termination of a pregnancy without primary surgical intervention and ‘EMA’ applies to these procedures in the first trimester.^14^ The recommended EMA regimen uses the anti-progestogen mifepristone in an oral dose of 200mg and the prostaglandin analogue misoprostol by the sublingual, vaginal or buccal route. A single dose of 800mcg misoprostol used 24-48 hours after mifepristone has been demonstrated to be highly effective up to 10 weeks’ gestation.^15^ Beyond 10 weeks additional doses of misoprostol are needed to maintain the same degree of effectiveness.^16^

Our dataset consisted of information on EMAs extracted directly from electronic service records and included fully de-identified patient clinical and demographic characteristics, as well as clinical outcomes. All data were extracted six weeks after the end of the study period to ensure the reporting of complications was as complete as possible.

Two cohorts were defined. The “traditional” cohort comprises all patients having an EMA between January 1^st^ and March 1st 2020, prior to service model change. All patients in this cohort received care using the traditional pathway that included an in-person assessment and an ultrasound scan. The “telemedicine-hybrid” cohort comprises all patients accessing an EMA between April 6^th^ and June 30^th^ 2020, in a two month period after the service model change at each provider. Patients in this cohort received care using no-test telemedicine if they had low risk of ectopic pregnancy and their self-reported last menstrual period (LMP) indicated a gestation of less than 10 weeks. Those who were not eligible for telemedicine had an in-person assessment with ultrasound. Providers followed organisation-specific evidence-based policies informed by the RCOG^9^ and associated decision aid^17^ (figures 1 and 2) and earlier RCOG and NICE guidelines for general abortion care including in-person medical abortion.^18 19^

**Figure 1.**
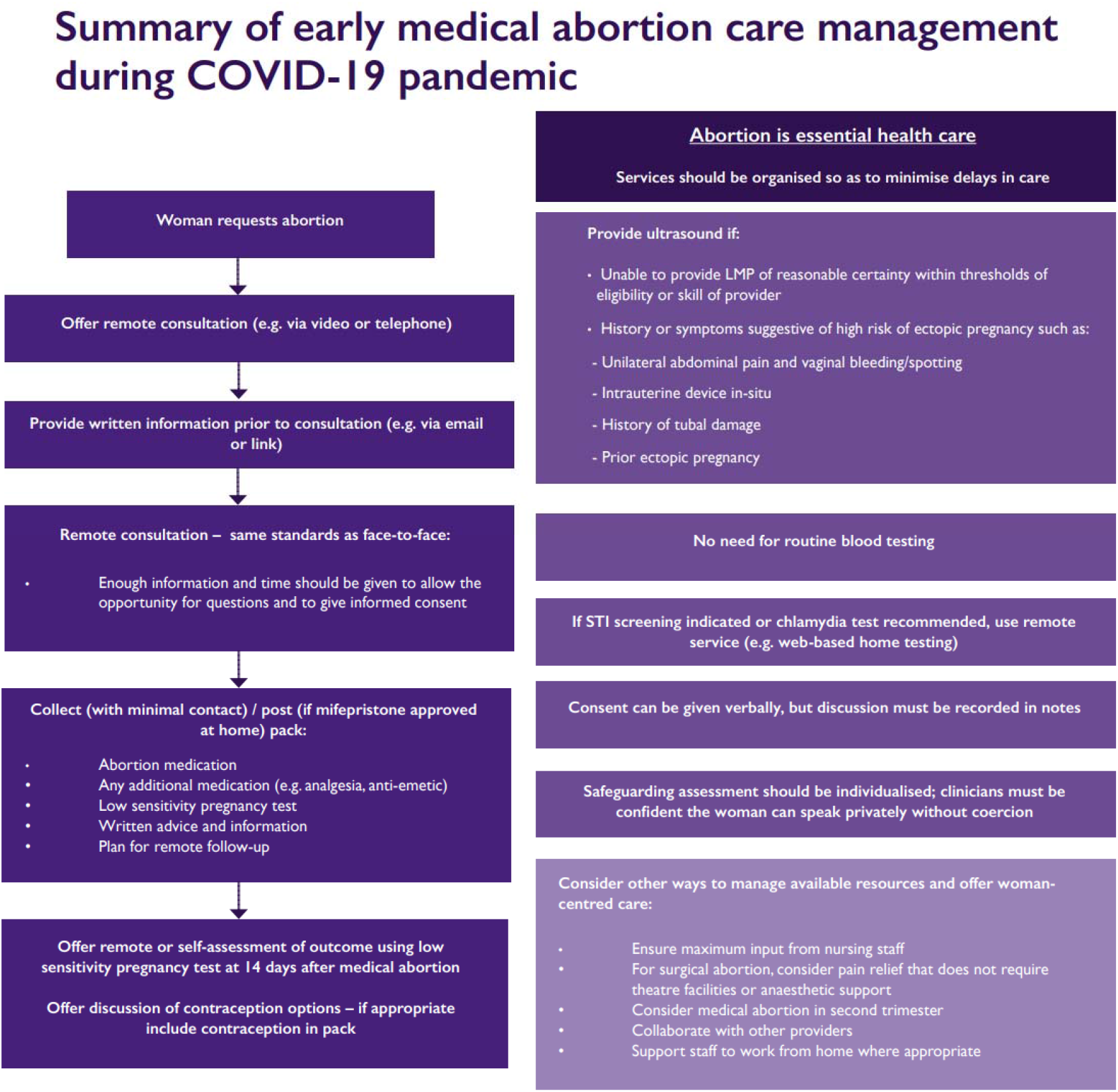
Summary of early medical abortion care management during COVID-19 pandemic (adapted from RCOG Coronavirus (COVID-19) infection and abortion care – information for healthcare professionals)^9^. [Note for editors – permission for reproduction obtained from RCOG] Image from page 4: 2020-07-31-coronavirus-covid-19-infection-and-abortion-care.pdf (rcog.org.uk) If preferred, can provide original files in original Word format

**Figure 2.**
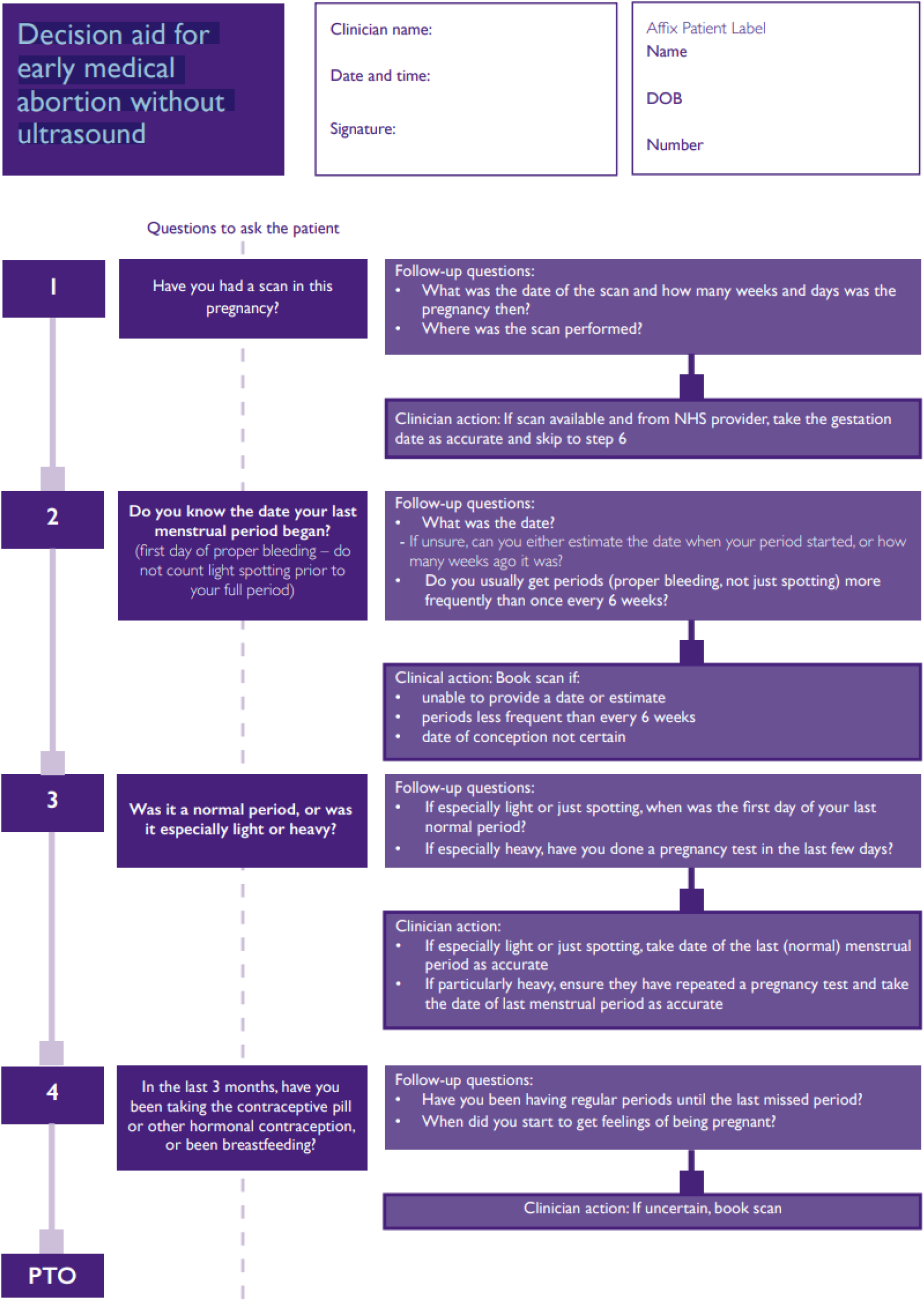
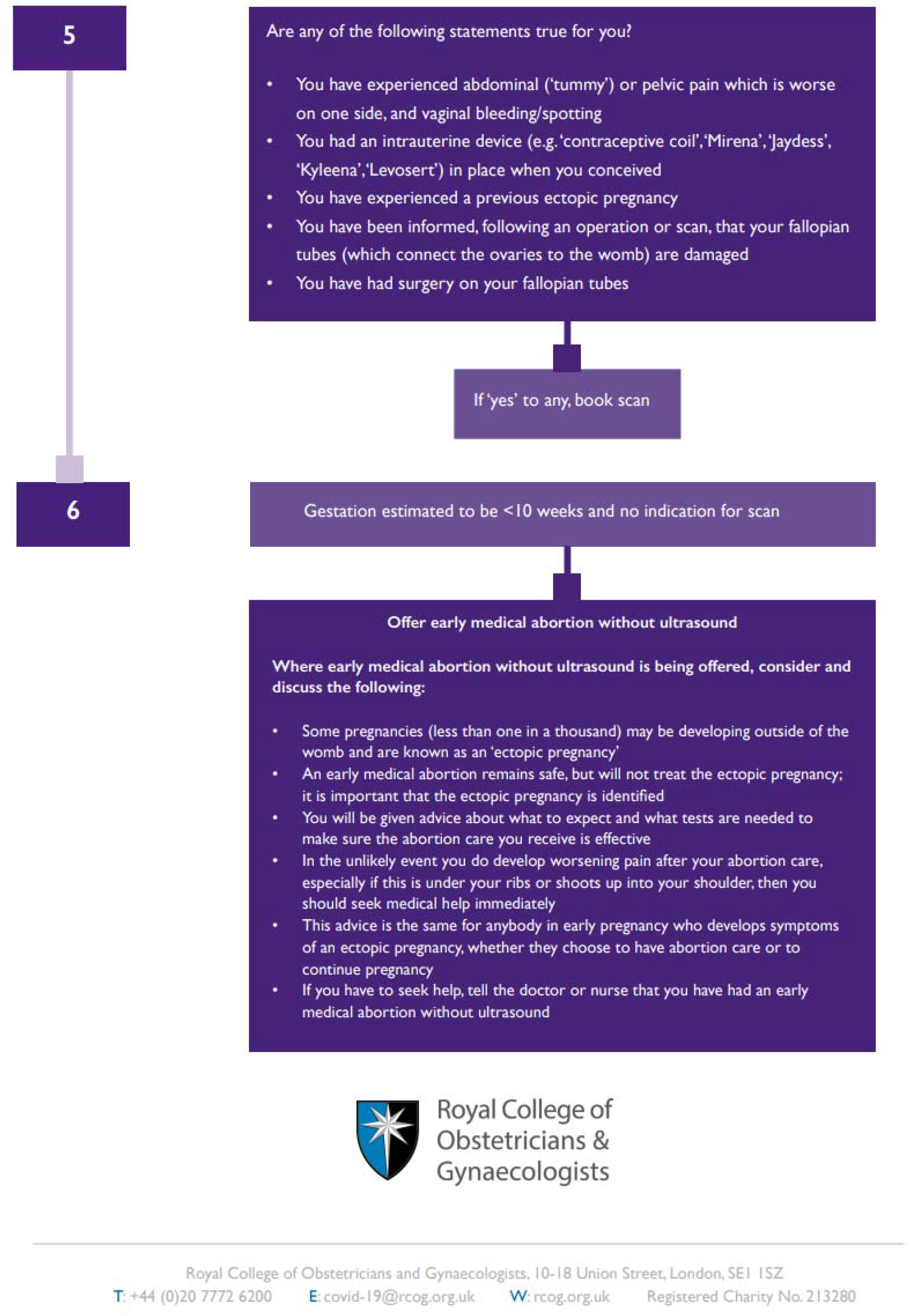
Decision aid for early medical abortion without ultrasound (adapted from RCOG Coronavirus (COVID-19) infection and abortion care – information for healthcare professionals)^17^. [Note for editors – permission for reproduction obtained from RCOG] Image from: 2020-06-04-decision-aid-for-early-medical-abortion-without-ultrasound.pdf (rcog.org.uk) If preferred, can provide original files in original Powerpoint format

In the traditional cohort, whilst some aspects of the pre-abortion consultation may have been carried out by phone, an in-person assessment with ultrasound scan and in-clinic administration of mifepristone were always required. In the telemedicine-hybrid cohort, we defined patients served by telemedicine as those who received medications for home use (either by mail or collection with minimal contact from a clinic) following a phone or video consultation and who did not receive an ultrasound scan or other tests.

We omitted the month of March 2020 because of the uncertainty and challenges posed by the emergence of COVID prior to the definitive service model change. The dates of the telemedicine-hybrid cohort take into account the roll-out of the telemedicine service across individual clinics within a provider network. For the analysis of ectopic pregnancies, we applied the timeframes stated above to all abortion consultations rather than all abortions provided because some patients with suspected ectopic pregnancies were referred to Early Pregnancy Assessment Units (EPAU) and did not proceed to an abortion.

## Outcome Measures

### Access

Waiting time and gestation at treatment were used to assess how the two models impacted access. Waiting time was defined as the interval from first contact with the abortion provider to when medication was dispensed (either in-person in clinic or posted). Gestation was recorded as that applicable on the date mifepristone was dispensed or provided. Analysis was of mean gestation and the proportion of abortions performed at ≤6 weeks’ gestation.

### Effectiveness

Effectiveness was defined as the proportion of medical abortions that were successful. Success was defined according to the MARE Guidelines as successful expulsion of an intrauterine pregnancy without need for surgical intervention.^14^ Sub-categories of unsuccessful abortion included: surgical management of continuing pregnancy, surgical evacuation of retained products of conception, and continuing pregnancy where the patient opted for continuation or the outcome is unknown.

### Safety

Safety was defined according to the proportions of medical abortions that involved one or more significant adverse events. We defined significant adverse events as haemorrhage requiring transfusion, significant infection requiring hospital admission, major surgery, and death. We also examined the incidence of ectopic pregnancy and when it was diagnosed in the care pathway. All patients referred for further diagnostics (e.g. serial βhCG monitoring) but who had no further treatment are included in the ectopic pregnancy group although a proportion of these will be pregnancy of unknown locations (PULs) that includes failed early intrauterine pregnancies. We analysed the proportion of cases where treatment was reported to have occurred at ≥10 weeks’ gestation in the telemedicine-hybrid cohort (this outcome does not apply to the traditional cohort since all gestations were determined by ultrasound scan).

### Acceptability

Given the constraints of delivering healthcare during COVID-19 it was not possible to follow up with all patients to capture patient-reported outcomes. Two of the providers (BPAS and MSUK) collected data on patient feedback during the study period. Patients were invited to provide feedback one to three weeks after treatment, either by telephone using a structured interview tool (MSUK)^20^ or using an online form (BPAS)^21^. We analysed data on questions reporting on satisfaction, future preference and privacy. Although the questions were similar, only the MSUK survey included questions on the privacy of teleconsultation. All contact was by a non-clinician who had not been involved in the patient’s care.

### Analysis

The primary analysis was to assess whether the telemedicine-hybrid care model (telemedicine with in-person provision when indicated) was non-inferior to the traditional service model (blanket in-person provision). Non-inferiority is established by comparing effectiveness and safety in the two cohorts under the null hypothesis that the rates of successful abortion and serious adverse events are not different.

We first compared patient demographic and clinical characteristics between the cohorts to assess the need to covariate-adjust our hypothesis tests for systematic differences in the two groups that might affect abortion outcomes. All hypothesis tests were covariate adjusted for patient age, race/ethnicity, gestational age, parity, and prior abortions using logistic regression and weighted risk differences.^22^

We evaluated effectiveness by testing the alternative hypothesis that the rate of successful medical abortion in the telemedicine-hybrid cohort is lower than in the traditional cohort using a covariate-adjusted test of difference in proportions. We also performed a chi-squared test to evaluate whether the distribution of unsuccessful abortion sub-categories differed between the cohorts. We evaluated safety by testing the alternative hypothesis that significant adverse events occurred at higher rates in the telemedicine-hybrid cohort compared to the traditional cohort using a covariate-adjusted hypothesis test for difference of proportions. We also evaluated whether the prevalence of ectopic pregnancies managed before EMA and after EMA were different between the traditional and telemedicine-hybrid cohorts using chi-squared difference of proportions tests.

The secondary analysis was to compare effectiveness and safety of medical abortion for patients who received fully remote no-test telemedicine vs. in-person care in the telemedicine-hybrid cohort, primarily to assess whether any differences between the cohort service models were driven by one particular group. We evaluated patient demographic and clinical characteristic differences between the two groups to assess the need to covariate-adjust our hypothesis tests for systematic differences and all hypothesis tests were covariate adjusted for patient age, race/ethnicity, gestational age, parity, and prior abortions using the framework described above. We note, however, that these cohorts are fundamentally different despite covariate adjustment, as patients in the telemedicine-hybrid cohort were selected into inperson treatment based on characteristics that would affect the outcome of their abortion. We performed covariate-adjusted hypothesis tests under the null hypothesis of equal effectiveness and equal rates of significant adverse events in the telemedicine vs. in-person groups.

All analyses were performed using R, version 3.6.2. Statistical significance was defined using an alpha level of 0.05.

### Ethics Approval

The study was reviewed by the Institutional Review Board (IRB) at the University of Texas at Austin and a determination was made that the research did not meet the criteria for human subjects research as defined in the Common Rule (45 CFR 46) or FDA Regulations (21 CFR 56). Each provider ensured compliance with their own internal ethics and governance systems. The principles of the STROBE statement, and the MARE supplement, were followed.^14^

## Results

In total, 52,142 medical abortions were provided during the study period; 22,158 in the traditional pathway and 29,984 in the telemedicine-hybrid cohort. Among those that took place in the telemedicine-hybrid cohort, 18,435 (61%) were provided entirely via telemedicine and 11,549 (39%) were provided inperson. Our sample represents 85% of the total number of medical abortions performed in England and Wales during the study period.^23^ The clinical and demographic characteristics of patients in the two cohorts are described in Table 1.

**Table 1:** Clinical and demographic characteristics in the traditional and telemedicine-hybrid cohorts (n=52,142)

| Patient Characteristic | Traditional<br>(n=22,158) | Telemedicine-hybrid<br>(n=29,984) | P-value |
| --- | --- | --- | --- |
| **Mean gestational age in weeks (s.d.)** | 6.4 (1.3) | 6.0 (1.4) | <0.001 |
| **Gestational age at treatment** |  |  |  |
| 6 weeks and under | 5,582 (25.2) | 11,947 (39.8) | <0.001 |
| Over 6 weeks | 16,576 (74.8) | 18,037 (60.2) |  |
| **Mean age in years (s.d.)** | 27.8 (6.6) | 28.5 (6.7) | <0.001 |
| **Ethnicity** |  |  |  |
| Asian | 2,038 (9.2) | 2,652 (8.8) | <0.001 |
| Black | 1,656 (7.5) | 2,282 (7.6) |  |
| Multiracial | 1,004 (4.5) | 1,361 (4.5) |  |
| White | 15,840 (71.5) | 20,910 (69.7) |  |
| Other | 489 (2.2) | 638 (2.1) |  |
| Unknown | 1,131 (5.1) | 2,141 (7.1) |  |
| **Previous abortions** |  |  |  |
| 0 | 13,098 (59.1) | 16,741 (55.8) | <0.001 |
| 1+ | 9,060 (40.9) | 13,243 (44.2) |  |
| **Parity** |  |  |  |
| 0 | 10,133 (45.7) | 11,741 (39.2) | <0.001 |
| 1+ | 12,025 (54.3) | 18,243 (60.8) |  |
| **Mean waiting time in days (s.d.)** | 10.7 (19.9) | 6.5 (13.5) | <0.001 |

### Access

Mean waiting time to treatment declined from 10.7 days (SD 19.9) in the traditional pathway to 6.5 days (SD 13.5) in the telemedicine-hybrid cohort (p<0.001). Mean gestational age at treatment also declined in the telemedicine-hybrid cohort resulting in 40% of abortions performed at 6 weeks’ gestation or less vs. 25% in the traditional pathway (p<0.001).

### Effectiveness

Rates of successful medical abortion were high under both service delivery models (Table 2): 98.2% in the traditional cohort vs. 98.8% in the telemedicine-hybrid cohort. We found no evidence of a lower success rate with the telemedicine-hybrid pathway (p=1.0). The distribution of the different sub-categories of unsuccessful medical abortion did not differ between the cohorts (p=0.268).

**Table 2:** Comparison of effectiveness of medical abortions conducted in the traditional and telemedicine-hybrid cohorts (n=52,142)

| Outcome | Traditional<br>n=22,158 | Telemedicine-hybrid<br>n=29,984 | P-value |
| --- | --- | --- | --- |
| **Successful medical abortion** | **21,769 (98.2)** | **29,618 (98.8)** | **1.0** |
| **Unsuccessful medical abortion** | **389 (1.8)** | **366 (1.2)** |  |
| Continuing pregnancy treated with surgical management | 161 (0.7) | 150 (0.5) | 0.268 |
| Continuing pregnancy opted to continue or unknown outcome | 3 (0.01) | 8 (0.03) |  |
| Retained products treated with surgical management (ERPC) | 225 (1.0) | 208 (0.7) |  |

Note: As explained in the methods section, the p-value for successful medical abortion is the co-variate adjusted p-value (i.e. all differences in patient clinical and demographic characteristics, including gestational age, are controlled for) and was calculated using a hypothesis test where the null hypothesis is that the traditional cohort has the same effectiveness rate as the telemedicine-hybrid cohort and the alternative hypothesis is that the traditional cohort has a higher effectiveness rate than the telemedicine-hybrid cohort. The p-value for unsuccessful medication abortion is for the chi-squared test of whether the distribution of types of failure differ between the cohorts.

### Safety

Significant adverse events in both cohorts were rare (Table 3). Haemorrhage requiring transfusion was reported in 8 (0.04%) cases in the traditional cohort and 7 (0.02%) cases in the telemedicine-hybrid cohort. No cases of significant infection requiring hospital admission, major surgery or death were reported. We found no evidence that significant adverse events were higher in the telemedicine-hybrid cohort (p=0.557).

**Table 3:**
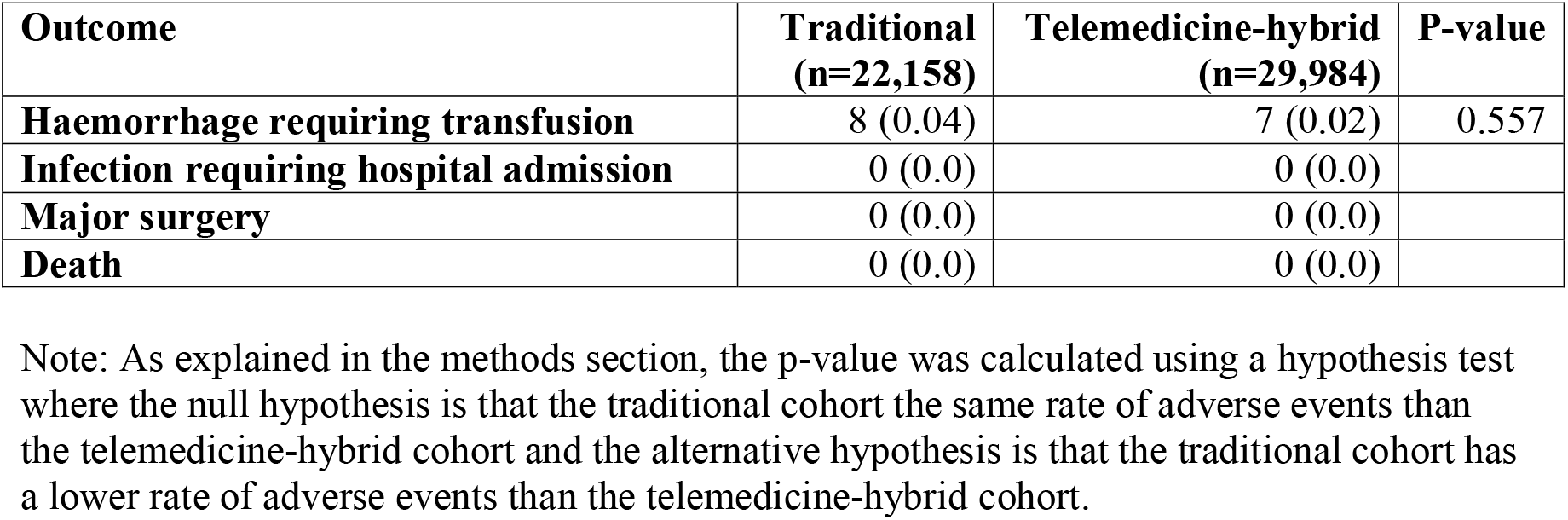
Comparison of significant adverse events following medical abortions conducted in the traditional and telemedicine-hybrid cohorts (n=52,142)

The overall incidence of ectopic pregnancy was equivalent in both cohorts, 39 (0.2%) in the traditional pathway and 49 (0.2%) in the telemedicine pathway, p=0.796 (Table 4). The proportions managed after EMA were not significantly different between the cohorts (0.01% in the traditional pathway and 0.03% in the telemedicine-hybrid pathway, p=0.123). There were 11 cases (0.04%) in the telemedicine-hybrid cohort where the gestational age after abortion was reported as being greater than the expected 10 weeks. In all these cases, the medical abortion was completed at home without additional medical complications.

**Table 4:** Significant outcomes among patients presenting for medical abortion in the traditional and telemedicine-hybrid cohorts (n=52,218)

| Outcome | Traditional<br>(n=22,197) | Telemedicine-hybrid<br>(n=30,021) | P-value |
| --- | --- | --- | --- |
| Ectopic Managed Pre-Treatment | 37 (0.17) | 39 (0.13) | 0.796 |
| Ectopic Managed Post-Treatment | 2 (0.01) | 10 (0.03) | 0.123 |
| Gestational age later than expected* | 0 (0.0) | 11 (0.04) | N/A |

Note: The column Ns include patients who presented for an EMA but did not receive one because their ectopic pregnancy was identified pre-treatment.
\*Note: The column Ns for the gestational age later than expected category are the same as those in Table 3, (all EMAs performed in the two cohorts, i.e. N = 52,142).

### Acceptability

Patient-reported outcome data was available from 2453 respondents. 96% were “satisfied” or “very satisfied” with their care, or rated their experience as “good” or “very good”. 80% reported that they would choose telemedicine in the future or that it was their preferred option, with 13% choosing in-person care and the remainder being unsure. No patient reported that they were unable to consult in private using teleconsultation (n=1243).

### Secondary Outcome Measures

Comparison of clinical outcomes for the telemedicine vs. in-person groups in the telemedicine-hybrid cohort are shown in Tables S1-S3. Rates of successful medical abortion were higher in the telemedicine group (99.2% vs 98.1%, p<0.001), and rates of significant adverse outcomes were not significantly different between the two groups, 3 (0.02%) for telemedicine vs. 4 (0.03%) for in-person, p=0.532.

## Discussion

### Principal Findings

We found that no-test telemedicine without routine ultrasound for medical abortion up to 10 weeks’ gestation is an effective, safe and acceptable service model. Clinical outcomes with telemedicine are equivalent to in-person care and access to abortion care is better, with both waiting times and gestational age at the time of abortion significantly reduced. Further evidence that the new telemedicine-hybrid model improved access comes from a study showing that the rate of women seeking abortion medication outside the formal healthcare setting reduced significantly in the UK following implementation of telemedicine.^24^ The most likely explanation is easier access as the opposite effect was seen in several other European countries who made no such provision. The implication is that those previously too vulnerable to attend in-person have been able to access care through telemedicine, potentially benefiting from the safeguarding, counselling and contraceptive services provided by regulated providers.

Our study confirms previous literature that medical abortion is safe and effective with low rates of significant complications.^25^ The slight increase in effectiveness we observed in the group that received telemedicine – even after controlling for lower average gestational age compared to the in-person group – may be due to the ability of patients to better control the timing at which they took the medication.

The telemedicine-hybrid model resulted in very low rates of undiagnosed ectopic pregnancy and later than expected gestations. Although the rate of ectopic pregnancy in the general population in the UK and USA is reported as 1-2%,^26 27^ the rate reported among patients having an abortion is 10 times lower,^28^ which is consistent with our findings. Ultrasound is not used to screen for ectopic pregnancy in the general population – it is only used where signs and symptoms suggest a need.^27^ Routine screening of symptom-free women is associated with a high false positive rate when the prevalence of ectopic pregnancy is low, as is the case in women seeking abortion, and therefore it is unlikely there would be significant benefits.^29^ There is no clinical justification for maintaining this inconsistency in care between women wishing to continue their pregnancies and those choosing EMA.^30 31^

However, given that over 200,000 people access abortion care each year in the UK alone, some will inevitably have an asymptomatic ectopic pregnancy and so will proceed with having mifepristone and misoprostol either through telemedicine or after a false negative scan. The essential issue for safety is that these are detected prior to causing harm rather than prior to beginning medical abortion treatment; treatment with mifepristone and misoprostol in itself will have no effect on an underlying ectopic pregnancy. Indeed, the reduction in waiting times afforded by the telemedicine model may facilitate earlier detection than traditional pathways where women present later, or are sent away to give additional time to visualise an intrauterine pregnancy on scan. Proceeding with early medical abortion without a scan may permit earlier diagnosis of a developing ectopic pregnancy owing to increased surveillance and index of suspicion, for example where there is minimal bleeding after misoprostol.^9 18^

The proportion of cases where gestational age was later than expected based on LMP was low, as might have been expected given the evidence that women can determine the gestational age of their pregnancy with reasonable accuracy by LMP alone.^31^ Nevertheless inadvertent treatment of gestations over 10 weeks is inevitable and, consistent with our findings, the consequences for most are unlikely to be medically significant.^32^ The 10 weeks’ gestation limit in the English government’s approval order is arbitrary, and not based on evidence of safety or effectiveness. The Scottish government did not stipulate a limit, leaving the decision to the discretion of the clinician in consultation with their patient. Moreover, the reported success of self-managed abortions at >12–24 weeks’ gestation is 93%, with safety similar to that expected in earlier gestations.^33^

### Strengths and Weaknesses

While the study is not a clinical trial and we were not able to control the assignment of patients to treatment groups to conduct a direct comparison of fully remote telemedicine with in-person care, the policy changes precipitated by COVID-19 created a rare opportunity to conduct an evaluation of a major change in service delivery model with a very large sample size. We were able to evaluate the outcomes of both the telemedicine-hybrid and traditional in-person services as they operate in the real world, and we were able to adjust for key covariates. A key strength of the study is the generalisability of our findings given that our sample included 85% of all medical abortions provided in England and Wales during the study period.

The main limitation of this study is that we were unable to actively follow-up patients postabortion and therefore only significant adverse events can be reported with confidence. There is a potential gap in the consistency of reporting incidents, due to some complications not meeting the threshold of serious incidents, multiple routes of entry into the NHS and informal communication between the NHS and abortion providers. Although it is possible that some patients presented to other providers and a significant adverse event was not reported in our dataset, the risk management and reporting systems within the NHS are well defined, with serious incidents being routinely shared with regulators who would expect providers to ensure actions had been taken to mitigate risks to patients in the future. More importantly, there is no reason to suspect that any under-reporting that did occur would be more likely in either cohort so as to introduce bias. The governing body of the NHS in England alerted all commissioners of the need to report incidents relating to telemedicine and there were review meetings of key stakeholders to ensure compliance. We consulted with regulators and national agencies to ensure that we accounted for reports made directly to them. The independent regulator of all health and social care services in England, the Care Quality Commission (CQC), confirmed that all cases reported directly to them through various routes, for example, statutory notifications and the central NHS database of patient safety incident reports (the National Reporting and Learning System (NRLS) and the Strategic Executive Information System (StEIS)), were known to the providers.

### Implications for Clinicians and Policymakers

This large study of 52,145 medical abortions demonstrates that incorporating no-test telemedicine into the care pathway is not inferior to the traditional pathway where all patients are seen in person and have an ultrasound scan. There are advantages - waiting times and gestation at abortion are reduced and it is highly rated by patients. There was no evidence of worse outcomes in failure rate, haemorrhage, need for surgery or in failure to detect ectopic pregnancy. In the 0.04% of cases where the abortion appeared to have been provided at over 10 weeks’ gestation, these were all completed at home without additional medical complications. Given the advantages of improving access to care, especially in vulnerable groups and in resource-poor healthcare systems or where patients often have to fund their own care, the evidence is compelling that no-test telemedicine should become routine in the provision of abortion care.

## Data Availability

The collated datasets, which include participant data with anonymised identifiers, are held by AA at the University of Texas. Consideration will be given to sharing this with bone fide researchers on application. The original data resides with the co-authors' own institutions. Although the data is de-identified, some relate to very rare events and could therefore result in identification. Therefore data on complications, and data arising from clinical incident reports, will be subject to the same access restrictions as those of the organisation supplying it.

## Supplementary Tables to Accompany Text in the Results Section

**Table S1:**
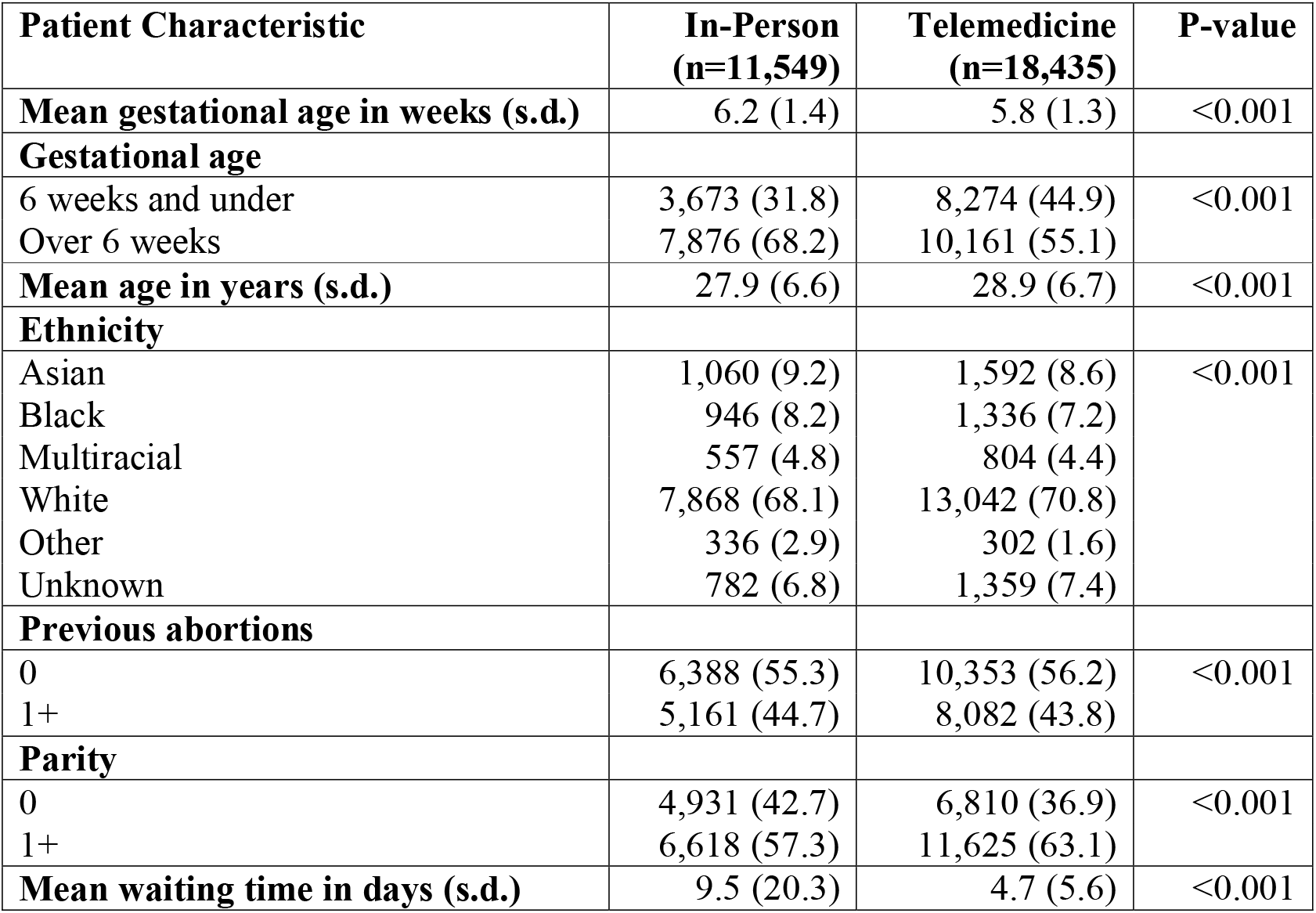
Patient clinical and demographic characteristics in the in-person vs. telemedicine groups for the telemedicine-hybrid cohort (n=29,984)

**Table S2:** Comparison of effectiveness of medical abortions conducted in the in-person vs. telemedicine groups for the telemedicine-hybrid cohort (n=29,984)

| Outcome | In-Person<br>n=11,549 | Telemedicine<br>N=18,435 | P-value |
| --- | --- | --- | --- |
| **Successful medical abortion** | **11,329 (98.1)** | **18,289 (99.2)** | **<0.001** |
| **Unsuccessful medical abortion** | **220 (1.9)** | **146 (0.8)** |  |
| Continuing pregnancy treated with surgical management | 133 (1.2) | 17 (0.09) | <0.001 |
| Continuing pregnancy opted to continue or unknown outcome | 1 (0.01) | 7 (0.04) |  |
| Retained products treated with surgical management (ERPC) | 86 (0.7) | 122 (0.7) |  |

Note: As explained in the Methods section, the p-value for successful medical abortion is the co-variate adjusted p-value (i.e. all differences in patient clinical and demographic characteristics, including gestational age, are controlled for) and was calculated using a hypothesis test where the null hypothesis is that the in-person has the same effectiveness rate as the telemed group and the alternative hypothesis is that the in-person group has a lower effectiveness rate than the telemedicine group. The p-value for unsuccessful medical abortion is for the chi-squared test of whether the distribution of types of failure differ between in-person and telemed groups.

**Table S3:** Comparison of significant adverse events following medical abortions in the in-person vs. telemedicine groups for the telemedicine-hybrid cohort (n=29,984)

| Outcome | In-Person<br>n=11,549 | Telemedicine<br>n=18,435 | P-value |
| --- | --- | --- | --- |
| Haemorrhage requiring transfusion | 4 (0.03) | 3 (0.02) | 0.532 |
| Infection requiring hospital admission | 0 (0.0) | 0 (0.0) |  |
| Major surgery | 0 (0.0) | 0 (0.0) |  |
| Death | 0 (0.0) | 0 (0.0) |  |

Note: As explained in the methods section, the p-value was calculated using a hypothesis test where the null hypothesis is that the in-person group has the same rate of adverse events than the telemedicine group and the alternative hypothesis is that the in-person group has a lower rate of adverse events than the telemedicine group.

## Acknowledgements

We would like to thank the Care Quality Commission (CQC), NHS England and NHS Improvement (NHSE & I), and the National Child Mortality Database (NCMD) for their advice and assistance in ensuring all complications reported within NHS systems have been included.

We are also enormously grateful to the many people who helped to ensure access to essential healthcare for women needing an abortion not only continued, but was improved during the COVID-19 pandemic. There are too many to list individually, but we would like to especially thank the following who not only helped to implement a complete service change within a few weeks, but also made this paper possible by ensuring the data was collected and accessible:

BPAS – Rebecca Blaylock, Steve Cheung, Pam Field, Stephen Franklin, Jeanette Taylor, Katherine Whitehouse

MSUK – Cat James, Andrew Lord, Kay Newey, Abigail Storan

NUPAS – Aaron Flaherty, Linda Leach, Leanne MacCaffrey

## Authors’ contributions & Declarations of Interest

All authors have completed the ICMJE uniform disclosure form at www.icmje.org/coi_disclosure.pdf and declare: no support from any organisation for the submitted work; no financial relationships with any organisations that might have an interest in the submitted work in the previous three years; no other relationships or activities that could appear to have influenced the submitted work beyond that described under the roles held by each individual below.

**Abigail Aiken** – Initial concept, developing study protocol, data analysis, writing first draft and subsequent revisions, verification of data. Overall responsibility for data analysis & principal investigator. Guarantor.

Member of the Council of the British Society of Abortion Care Providers (BSACP).

**Nabanita Ghosh** – Developing study protocol, revision of draft paper. Overall responsibility for conduct of study including data collection at NUPAS.

Nothing to disclose

**Patricia A. Lohr** – Initial concept, developing study protocol, writing first draft and subsequent revisions. Overall responsibility for conduct of study including data collection at BPAS.

Co-author / committee member of national guidelines cited in this paper from RCOG & NICE; member RCOG abortion taskforce; council member of British Society of Abortion Care Providers (BSACP); Medical Director of BPAS.

**Jonathan Lord** – Initial concept, developing study protocol, writing first draft and subsequent revisions, co-ordination and liaison. Overall responsibility for conduct of study including data collection at MSUK.

Co-author / committee member of national guidelines cited in this paper from RCOG & NICE; co-chair of RCOG abortion taskforce and British Society of Abortion Care Providers (BSACP); primary employment includes work as an abortion care provider as a consultant gynaecologist and medical director.

**Jennifer Starling** – Data analysis including statistical advice, writing first draft and subsequent revisions, verification of data.

Nothing to disclose

## Copyright/license for publication

The Corresponding Author has the right to grant on behalf of all authors and does grant on behalf of all authors, a worldwide licence to the Publishers and its licensees in perpetuity, in all forms, formats and media (whether known now or created in the future), to i) publish, reproduce, distribute, display and store the Contribution, ii) translate the Contribution into other languages, create adaptations, reprints, include within collections and create summaries, extracts and/or, abstracts of the Contribution, iii) create any other derivative work(s) based on the Contribution, iv) to exploit all subsidiary rights in the Contribution, v) the inclusion of electronic links from the Contribution to third party material where-ever it may be located; and, vi) licence any third party to do any or all of the above

## Transparency Statement

The authors affirm that the manuscript is an honest, accurate, and transparent account of the study being reported; that no important aspects of the study have been omitted; and that any discrepancies from the study as originally planned (and, if relevant, registered) have been explained.

